# Why Do Some Football Fans Experience More Anxiety? Insights from the 2022 FIFA World Cup

**DOI:** 10.64898/2026.01.16.26344288

**Authors:** Mohammed A. Alarabi, Nasser S. Alharbi, Fadi Aljamaan, Elshazaly Saeed, Shereen A. Dasuqi, Ibraheem Altamimi, Reem Alageel, Hadeel Alsulami, Yazan Chaiah, Amr Jamal, Khaled Saad, Fahad A. Bashiri, Khalid Alhasan, Ahmed S. BaHammam, Shuliweeh Alenazi, Mohamad-Hani Temsah

## Abstract

**Background:** Large sporting events, such as the FIFA World Cup, attract global attention and raise questions about psychological impact. Anxiety is among the most common mental health symptoms, but little is known about its predictors during international tournaments. This study explored predictors of anxiety during the FIFA 2022 World Cup.

**Methods:** This cross-sectional study included 1,331 respondents mostly residing in Saudi Arabia who completed an online survey during the FIFA 2022 World Cup (November–December 2022). The survey included sociodemographic variables, items related to the World Cup viewing habits, perceived effects of the tournament, and the Generalized Anxiety Disorder 7-item (GAD-7) scale.

**Results:** Although most respondents reported minimal anxiety symptoms (GAD-7 median score = 5, IQR = 6), 41.5% of the sample felt that watching the World Cup made them feel nervous. Females and younger adults reported significantly higher GAD-7 scores (*p* < 0.001). Employment status and method of watching the matches were not significant predictors. Regression modelling revealed that age, gender, and perceived sleep disruption predicted 9.1% of the variance in GAD-7 scores (*p* < 0.001). Younger age was the strongest predictor, while perceived sleep disturbance related to watching the matches predicted higher GAD-7 scores.

**Conclusions:** This was a population-based study of anxiety during the FIFA 2022 World Cup. Younger adults were more likely to report elevated anxiety, and sleep disruption was linked to higher GAD-7 scores. These findings highlight modifiable and non-modifiable predictors of anxiety and may inform public health strategies ahead of FIFA 2026.

## Introduction

The FIFA World Cup stands as one of the most watched and emotionally engaging sporting events worldwide. The 2022 World Cup in Qatar confirmed this unprecedented reach, with a record five billion supporters around the world engaging with the ground-breaking tournament across all media (1). This massive audience highlighted the event’s profound cultural and social impact. Such intense global engagement, however, also raises critical public health questions about the psychological impact of these emotionally charged competitions. While the athletic spectacle is celebrated, the potential effects on the mental well-being of its vast audience, from exhilaration to significant stress, warrant scientific investigation.

Anxiety disorders represent one of the most pervasive mental health challenges globally, significantly impairing psychological welfare and social functioning, particularly among adolescents and young adults. These disorders typically manifest between the ages of 10 and 24, leading to severe health losses and role impairments(2, 3). The literature indicates that the global incidence of anxiety disorders among those aged 10-24 years increased by 52% from 1990 to 2021. Females consistently show higher prevalence rates than males, and Disability-Adjusted Life Years (DALYs) have risen notably among 20-24-year-olds(4).

(5–9)Against this evidence of rising anxiety prevalence, large-scale international events like the FIFA World Cup present unique, population-level psychosocial stressors. While much research has focused on the physical health impacts of sporting events, such as their triggering effect on cardiovascular events (10), the psychological consequences remain less explored. For example, during the 2006 FIFA World Cup in Germany, the incidence of cardiac emergencies in the greater Munich area was 2.66 times higher on days when the German team played compared to a control period (11). This demonstrates the significant physiological impact of emotional stress associated with major sporting events. However, a parallel understanding of the mental health impact, specifically regarding anxiety symptoms during such tournaments, is lacking.

Therefore, this study aimed to explore the predictors of anxiety symptoms during the FIFA 2022 World Cup within a predominantly Saudi Arabian population. This research investigated sociodemographic factors, viewing habits, and perceived effects the tournament had on audience’s self-reported anxiety symptoms. Understanding the predictors of anxiety is crucial for developing targeted public health strategies to mitigate adverse mental health outcomes during future high-stakes worldwide events.

## Methods

### Study Design

This study adopted a cross-sectional survey design conducted during the FIFA World Cup tournament in Qatar (November–December 2022). The aim was to examine self-reported anxiety symptoms and their predictors among adults following the tournament. The structure of the design parallels the approach used in the prior FIFA–related studies, which also employed cross-sectional online surveys during the World Cup viewing period (12).

### Sampling, Participants, and Data Collection

Survey invitations were disseminated via popular social media platforms (WhatsApp, Twitter (now X), and Facebook) utilizing convenience sampling methods, similar to the approach used in the children’s sleep surveys (12, 13). Eligible participants were adults aged 18 years or older, residing primarily in Saudi Arabia or internationally. Participants completed the questionnaire voluntarily after providing electronic informed consent. No identifying data were collected, ensuring anonymity and confidentiality throughout the study period.

### Survey Instrument

The survey included three major components:

1. Sociodemographic characteristics (age, gender, education, occupation, and residence).
2. FIFA World Cup viewing habits, including preferred method of viewing the matches and perceived effects of the tournament.
3. Generalized Anxiety Disorder Scale (GAD-7), a validated 7-item screening tool assessing anxiety symptoms over the previous two weeks (14). The GAD-7 has demonstrated strong reliability and is widely used in epidemiological mental health research. Item scoring ranges from 0 (“not at all”) to 3 (“nearly every day”), yielding a total score from 0 to 21, with higher scores indicating more severe anxiety symptoms.

### Sample Size Considerations

A priori sample size estimation was based on an assumed 50% prevalence of anxiety-related symptoms, providing a conservative estimate to ensure adequate power. The required minimum sample size for a 95% confidence level and a 5% margin of error was 385 participants. (12)

### Ethical Considerations

The study protocol was reviewed and approved by the appropriate Institutional Review Board at King Saud University, Riyadh, Saudi Arabia (approval number: 19/0953/IRB). Participation was voluntary, and respondents could withdraw at any time without consequence. All data were stored securely and analyzed anonymously.

#### Data Analysis

All analyses were conducted using IBM SPSS Statistics for Windows (Version 21, IBM Corp, Armonk, NY). Descriptive statistics were used to report sample characteristics, with continuous variables reported as means (SD) or medians (IQR) as appropriate, and categorical variables reported as frequencies and percentages. Normality was tested using the Shapiro–Wilk test. Between-group differences were assessed using the independent samples t-test or Mann– Whitney U test for continuous variables and Chi-square test for categorical variables. Variables showing significant bivariate associations (*p* < 0.05) were entered into a multivariable linear regression model using the enter method.

## Results

### Demographics

The data of 1331 respondents were included in the analysis. The distribution of age and gender is presented in Table 1. The sample was slightly skewed toward female respondents (52.7%), and the largest age group was 18–24 years (33.0%). This indicates that younger adults, particularly women, were the most represented demographic in this survey.

**Table 1.** Distribution of Age Group and Gender Amongst Survey Respondents (N = 1331):

|  | Category | Frequency | Percent |
| --- | --- | --- | --- |
| **Age group (years)** | 18–24 | 439 | 33.0 |
|  | 25–34 | 335 | 25.2 |
|  | 35–44 | 291 | 21.9 |
|  | 45–54 | 170 | 12.8 |
|  | 55–64 | 81 | 6.1 |
|  | 65+ | 15 | 1.1 |
| **Gender** | Female | 702 | 52.7 |
|  | Male | 629 | 47.3 |
|  | Total | 1331 | 100.0 |

Regarding the highest level of education, the majority of respondents reported a university or postgraduate degree (n = 1076, 80.8%). High school was the second most common level (n = 199, 15.0%), followed by middle school (n = 24, 1.8%), primary school (n = 8, 0.6%), and other educational backgrounds (n = 24, 1.8%).

Employment status showed that 449 respondents (33.7%) were unemployed, while 882 (66.3%) reported holding jobs. Among the working respondents, 417 (31.3% of the total sample) were in regular employment, 295 (22.2%) worked in the healthcare sector, 63 (4.7%) reported freelance work, and 107 (8.0%) reported other forms of occupation.

With respect to the country of residence, we gathered 392 responses (29.5%) with the remainder of the sample not reporting this variable. Of the respondents who reported their country of residence, most of them (n = 361, 92.1%) reported living in Saudi Arabia. A smaller proportion (n = 31, 7.9%) resided outside Saudi Arabia. The most frequently reported countries were the United States (n = 8) and Kuwait (n = 7). Canada (n = 3) and Oman (n = 2) were also reported, while single responses were reported from Bahrain, Bangladesh, Egypt, Philippines, Qatar, Syria, Sudan, Yemen, and the United Arab Emirates.

### Anxiety Scores

The mean total score on the GAD-7 was 5.84 (SD = 5.06, 95% CI: 5.57–6.11), with a median of 5 and an observed range from 0 to 21 (possible range 0–21). The interquartile range was 6, indicating that half of the sample scored between 2 and 8. The distribution was positively skewed (skewness = 0.92), with a Shapiro–Wilk test confirming non-normality (*p* < 0.001). This suggests that while most respondents reported low anxiety symptoms, a smaller proportion reported higher scores.

Item-level analysis showed that the most frequently endorsed symptoms were “worrying too much about different things” and “feeling nervous, anxious, or on edge,” both with median scores of 1. In contrast, “restlessness” and “feeling afraid as if something awful might happen” had median scores of 0, indicating respondents reported these symptoms rarely or not at all (Table 2).

**Table 2.** Descriptive Statistics for GAD-7 Scores (N = 1331)

| Item | Mean (SD) | Median | IQR |
| --- | --- | --- | --- |
| 1. Feeling nervous, anxious, or on edge | 0.97 (0.91) | 1.00 | 1.00 |
| 2. Not being able to stop or control worrying | 0.83 (0.92) | 1.00 | 1.00 |
| 3. Worrying too much about different things | 1.08 (0.98) | 1.00 | 2.00 |
| 4. Trouble relaxing | 0.88 (0.89) | 1.00 | 1.00 |
| 5. Being so restless that it is hard to sit still | 0.59 (0.85) | 0.00 | 1.00 |
| 6. Becoming easily annoyed or irritable | 0.92 (0.94) | 1.00 | 1.00 |
| 7. Feeling afraid as if something awful might happen | 0.58 (0.88) | 0.00 | 1.00 |
| **Total score** | **5.84 (5.06)** | **5.00** | **6.00** |

### Perception of FIFA World Cup

With respect to the psychological impact of the FIFA tournament, 553 respondents (41.5%) reported that watching the World Cup made them feel nervous, while 587 (44.1%) indicated no effect on their stress or relaxation, and 191 (14.4%) reported feeling more relaxed. Regarding sleep, 129 respondents (9.7%) stated that their sleep hours were greatly affected by watching the matches, 315 (23.7%) reported being affected a little, and the majority (n = 887, 66.6%) reported no change. Viewing habits during the World Cup varied across the sample. More than half of respondents (52.6%) reported watching matches live on television or mobile devices at home, while nearly one quarter (24.2%) watched live in public venues such as sports cafés. Smaller proportions followed match results live online (5.6%) or checked results after the matches had ended (8.6%). A further 9.0% indicated that they did not follow the World Cup, and 3.5% did not specify how they followed the matches.

### Predictors of Anxiety During The World Cup

When comparing anxiety scores across age groups, a significant difference was observed in GAD-7 total scores (Kruskal–Wallis χ^2^ (2) = 61.43, *p* < 0.001). Younger respondents reported higher levels of anxiety, with the highest mean rank among those aged 18–24 years, followed by 25–34 years, and 35–44 years. The lowest ranks were observed in older age groups, particularly those aged 55–64 years and 65 years and above. A median split confirmed this trend with 54.9% of respondents aged 18–24 years scoring above the sample median of 5, compared with only 24.7% of those aged 55–64 years. Furthermore, when comparing anxiety scores by gender, female respondents reported significantly higher levels of anxiety than males (Mann–Whitney U = 178,198.5, Z = –6.11, *p* < 0.001). This indicated that women were more frequently represented among those with elevated GAD-7 scores.

No significant differences in GAD-7 scores were observed across levels of highest education (Kruskal–Wallis χ^2^ (2) = 7.48, p = 0.112). When examining anxiety scores by employment status, a significant overall difference was detected across groups (Kruskal–Wallis χ^2^ (2) = 10.80, *p* = 0.029). Unemployed respondents had the highest mean rank, while the lowest ranks were observed among freelancers. A median split analysis, however, did not confirm a statistically significant association between employment status and being above the sample median of 5 (χ^2^ (2) = 6.53, p = 0.163).

No significant differences in GAD-7 scores were found based on how respondents watched the World Cup matches (Kruskal–Wallis χ^2^ (2) = 4.12, *p* = 0.390). A median split analysis confirmed the absence of association between viewing patterns and scoring above the sample median of 5 (χ^2^ (2) = 2.67, *p* = 0.615). On the other hand, reported changes in sleep due to watching the World Cup were significantly associated with anxiety scores (Kruskal–Wallis χ^2^ (2) = 10.31, *p* = 0.006). Respondents who reported that their sleep was greatly affected had the highest mean rank, followed by those who reported being slightly affected, while those with no reported change had the lowest rank. A median split confirmed this pattern with 55.8% of those with markedly affected sleep and 50.8% of those slightly affected scoring above the sample median of 5, compared with 43.3% of those who reported no change in sleep.

Because residuals from the initial linear regression model on raw GAD-7 scores were not normally distributed, the dependent variable was log-transformed to improve model fit. As detailed in Table 3, the regression model including age, gender, perceived impact of the World Cup on sleep, and employment status was statistically significant (F (4,1326) = 33.14, *p* < 0.001), explaining 9.1% of the variance in anxiety scores (adjusted R^2^ = 0.088). Female gender (B = 0.145, *p* < 0.001), age group (B = −0.068, *p* < 0.001), and reporting that sleep was affected by watching the World Cup (B = 0.062, *p* < 0.001) were significant predictors of higher anxiety scores, whereas unemployment status was not significantly associated (*p* = 0.909).

**Table 3.**
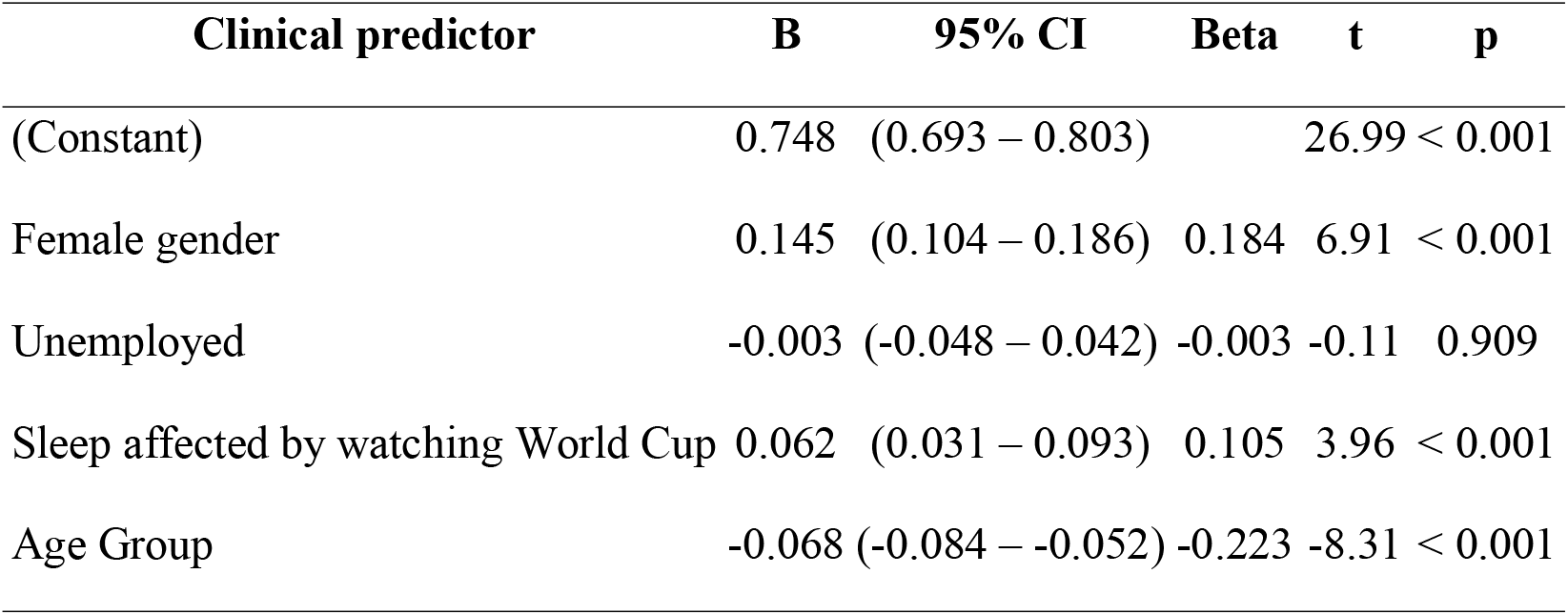
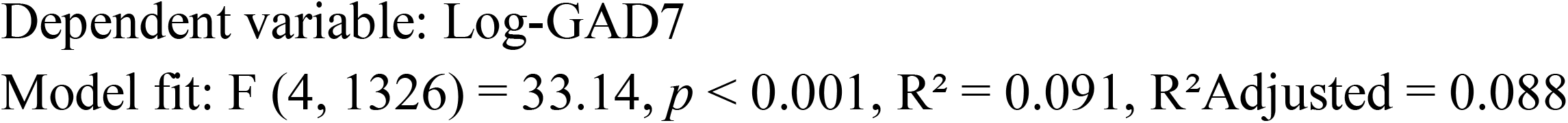
Linear Regression Model Predicting Log-GAD7 Scores:

## Discussion

This population-based study provides important insights into the predictors of anxiety symptoms during the FIFA 2022 World Cup. Our findings indicate that even within the context of a celebratory global event, a proportion of individuals experience heightened anxiety, with specific demographic and behavioral factors acting as key predictors. The most compelling findings of our study relate to non-modifiable demographic predictors. We found that younger age was the strongest predictor of higher GAD-7 scores, with anxiety decreasing progressively with older age groups. This is evidently illustrated by our data showing that 54.9% of respondents aged 18-24 scored above the sample median GAD-7 score of 5, compared to only 24.7% of those aged 55-64. This finding is highly consistent with the broader epidemiological literature on anxiety. This trend mirrors a broader, concerning pattern observed in the general population. A national survey in the United States documented a steep rise in anxiety among young adults, with the prevalence among those aged 18–25 years old more than doubling from 7.97% in 2008 to 14.66% in 2018, a significantly more rapid increase than in older age groups (15). This is further corroborated by a large, recent retrospective study in the US, which showed that the one-year prevalence of GAD more than tripled, rising from 2.1% in 2012 to 7.4% in 2022 (16). Anxiety disorders often have a median age of onset of 11 years, and their presence at an earlier developmental stage increases the likelihood of persistence into later life (6, 17). The heightened representation of young adults in our study with elevated anxiety could be linked to greater emotional investment in the tournament’s outcomes, social pressures related to national identity, or the use of social media, which can amplify stress during live events.

Furthermore, our results align with a vast body of evidence indicating that females are disproportionately affected by anxiety disorders (4, 7, 9, 17). We found that female respondents reported significantly higher anxiety levels than males. Globally, the prevalence of anxiety is approximately twice as high among women as among men (6, 18). (19) Our regression model confirmed female gender as a significant predictor. This gender disparity may be influenced by a combination of biological susceptibility, sociocultural factors, and differences in symptom reporting.

Beyond demographics, a key modifiable factor emerged as a significant predictor: perceived sleep disruption. Our regression model indicated that reporting sleep affected by watching the World Cup was significantly associated with higher log-GAD7 scores. This finding is greatly supported by the established and profoundly bidirectional relationship between sleep and anxiety, where anxiety interferes with sleep quality, while insufficient or disturbed sleep intensifies anxiety symptoms. This vicious cycle is especially pronounced in GAD, where up to 90% of individuals report insomnia symptoms, making it not just a symptom but potentially a diagnostic hallmark (9). Patients often describe distressing thoughts, heightened vigilance, and somatic hyperarousal that prevent them from falling asleep or cause them to wake frequently during the night. While insomnia has traditionally been seen as a consequence of anxiety, recent research suggests a bidirectional relationship, where insomnia also exacerbates, sustains, and may even precede the development of anxiety symptoms. In our sample, 55.8% of those who reported their sleep was “greatly affected” by the World Cup scored above the median anxiety score, compared to 43.3% of those with no sleep change. The interplay is complex and mutually reinforcing; the excessive cognitive arousal and worry characteristic of anxiety can interfere with sleep onset and maintenance, while sleep disruption can impair emotional regulation, thereby heightening vulnerability to anxiety (9).

(20)It is noteworthy that the method of watching matches was not a significant predictor of anxiety in our study. This suggests that the psychological engagement with the event, rather than the specific medium of consumption, is more critical in influencing anxiety levels. However, this finding does not preclude the possibility that pre-existing behavioral patterns linked to screen time and anxiety are at play. For instance, studies have shown that watching television for more than 120 minutes per day is associated with poorer sleep quality, a relationship that may be mediated by anxiety and other health conditions (21). In adolescents, both increased TV viewing and consumption of ultra-processed foods are independently associated with higher odds for anxiety-induced sleep disturbance (22). While our study did not find a direct link between viewing method and anxiety, the content and emotional intensity of what is being watched, such as a high-stakes penalty shootout, are likely stronger triggers than the platform itself.

Finally, our regression model, which included age, gender, sleep disruption, and employment status, was statistically significant but explained a relatively modest 9.1% of the variance in anxiety scores. This indicates that while we identified important predictors, a substantial portion of the variance is attributable to other factors not measured in our study. These could include pre-existing mental health conditions, genetic predispositions (6, 7), specific match outcomes, and the use of substances (7, 8).

## Limitations and Strengths

This study has several limitations. The cross-sectional design precludes any causal inferences. For instance, we cannot determine whether sleep disruption caused higher anxiety, or whether more anxious individuals were more likely to experience or report sleep problems, or both. The sample, while large (N=1,331), was predominantly composed of highly educated individuals (80.8% held a university or postgraduate degree) and resided mainly in Saudi Arabia (92.1% of reported country of residence), which may limit the generalizability of the findings to other cultural, educational, or geographic contexts. The reliance on self-reported measures for anxiety, sleep disruption, and viewing habits is subject to recall and social desirability biases.

Despite these limitations, this study possesses notable strengths. It is the first to quantitatively investigate anxiety predictors during a FIFA World Cup using a validated instrument (GAD-7) in a large population-based sample. The focus on a well-defined, real-world stressor provides high ecological validity. The examination of both modifiable and non-modifiable factors offers actionable insights into public health planning. Finally, our findings, which focused on psychological symptoms, complement the existing body of work on the physical health impact of major sporting events.

## Data Availability

All data produced in the present study are available upon reasonable request to the corresponding author

## Acknowledgement

The authors thank all participants in this survey, including parents, care givers and data collectors. While preparing this manuscript, the authors used ChatGPT-5 to refine the language and grammar, then the authors reviewed and edited the content as needed and they take full responsibility for the content of the submitted manuscript. The authors are grateful to the SABIC Research Chair.

## Conclusion

This study identified younger age, female gender, and perceived sleep disruption as significant predictors of anxiety symptoms during the 2022 FIFA World Cup. The findings underscore that mega-sporting events are powerful public health phenomena with measurable psychological consequences, mirroring the pattern of physical health risks previously established. While age and gender represent non-modifiable risk factors, sleep disruption associated with watching football matches emerged as a critical, modifiable target for intervention. As the world prepares for future sporting spectacles like the FIFA 2026 World Cup, public health strategies should extend beyond physical health preparedness to include mental well-being. Proactive health messaging could promote sleep hygiene and emotional self-care strategies, particularly targeting younger and female demographics, to help mitigate the psychological impact and foster a healthier, more resilient engagement with these global events.

## References

1. FIFA, Inside. Reports detail record global audience figures and positive sustainability outcomes from FIFA World Cup Qatar 2022™. 2024. Available from: https://inside.fifa.com/organisation/news/reports-detail-record-global-audience-figures-and-positive-sustainability-outcomes-fifa-world-cup-qatar.

2. Konnopka A, König H. Economic Burden of Anxiety Disorders: A Systematic Review and Meta-Analysis. Pharmacoeconomics. 2020 Jan;38(1):25–37. eng. doi:10.1007/s40273-019-00849-7. Cited in: Pubmed; PMID 31646432.

3. Castaldelli-Maia JM, Bhugra D. Analysis of global prevalence of mental and substance use disorders within countries: focus on sociodemographic characteristics and income levels. Int Rev Psychiatry. 2022 Feb;34(1):6–15. eng. Epub 20220219. doi:10.1080/09540261.2022.2040450. Cited in: Pubmed; PMID 35584016.

4. Bie F, Yan X, Xing J, Wang L, Xu Y, Wang G, Wang Q, Guo J, Qiao J, Rao Z. Rising global burden of anxiety disorders among adolescents and young adults: trends, risk factors, and the impact of socioeconomic disparities and COVID-19 from 1990 to 2021. Front Psychiatry. 2024;15:1489427. eng. The authors declare that the research was conducted in the absence of any commercial or financial relationships that could be construed as a potential conflict of interest. Epub 20241126. doi:10.3389/fpsyt.2024.1489427. Cited in: Pubmed; PMID 39691785.

5. Penninx BW, Pine DS, Holmes EA, Reif A. Anxiety disorders. Lancet. 2021 Mar 6;397(10277):914–927. eng. Epub 20210211. doi:10.1016/s0140-6736(21)00359-7. Cited in: Pubmed; PMID 33581801.

6. Munir S, Takov V. Generalized Anxiety Disorder. StatPearls. Treasure Island (FL): StatPearls Publishing Copyright © 2025, StatPearls Publishing LLC.; 2025. eng.

7. Mishra AK, Varma AR. A Comprehensive Review of the Generalized Anxiety Disorder. Cureus. 2023 Sep;15(9):e46115. eng. The authors have declared that no competing interests exist. Epub 20230928. doi:10.7759/cureus.46115. Cited in: Pubmed; PMID 37900518.

8. Zimmermann M, Chong AK, Vechiu C, Papa A. Modifiable risk and protective factors for anxiety disorders among adults: A systematic review. Psychiatry Res. 2020 Mar;285:112705. eng. Declaration of Competing Interest None. Epub 20191204. doi:10.1016/j.psychres.2019.112705. Cited in: Pubmed; PMID 31839417.

9. Xue Y, Wang WD, Liu YJ, Wang J, Walters AS. Sleep disturbances in generalized anxiety Disorder: The central role of insomnia. Sleep Med. 2025 Aug;132:106545. eng. Declaration of competing interest The authors declare that they have no known competing financial interests or personal relationships that could have appeared to influence the work reported in this paper. Epub 20250429. doi:10.1016/j.sleep.2025.106545. Cited in: Pubmed; PMID 40318600.

10. Mohammad MA, Karlsson S, Haddad J, Cederberg B, Jernberg T, Lindahl B, Fröbert O, Koul S, Erlinge D. Christmas, national holidays, sport events, and time factors as triggers of acute myocardial infarction: SWEDEHEART observational study 1998-2013. Bmj. 2018 Dec 12;363:k4811. eng. Competing interests: All authors have completed the ICMJE uniform disclosure form at https://www.icmje.org/coi_disclosure.pdf xand declare: no support from any organisation for the submitted work; no financial relationships with any organisations that might have an interest in the submitted work in the previous three years; no other relationships or activities that could appear to have influenced the submitted work. Epub 20181212. doi:10.1136/bmj.k4811. Cited in: Pubmed; PMID 30541902.

11. Wilbert-Lampen U, Leistner D, Greven S, Pohl T, Sper S, Völker C, Güthlin D, Plasse A, Knez A, Küchenhoff H, Steinbeck G. Cardiovascular events during World Cup soccer. N Engl J Med. 2008 Jan 31;358(5):475–83. eng. doi:10.1056/NEJMoa0707427. Cited in: Pubmed; PMID 18234752.

12. Temsah M-H, Aljamaan F, Altamimi I, Alageel R, Alsulami H, Dasuqi SA, Albabtain MA, Jamal A, Alenazi S, Alarabi M. Impact of FIFA World Cup 2022 on Children’s Sleep Patterns: An International Survey. medRxiv. 2025:2025–09.

13. Aljamaan F, Alhuzaimi A, Dasuqi SA, Alharbi N, Altamimi I, Alageel R, Alsulami H, Jamal A, Alenezi S, Alarabi M. Parental Perception of Children Sleep Pattern Changes During FIFA 2022. medRxiv. 2025:2025–12.

14. AlHadi AN, AlAteeq DA, Al-Sharif E, Bawazeer HM, Alanazi H, AlShomrani AT, Shuqdar RM, AlOwaybil R. An arabic translation, reliability, and validation of Patient Health Questionnaire in a Saudi sample. Ann Gen Psychiatry. 2017;16:32. eng. Epub 20170906. doi:10.1186/s12991-017-0155-1. Cited in: Pubmed; PMID 28878812.

15. Goodwin RD, Weinberger AH, Kim JH, Wu M, Galea S. Trends in anxiety among adults in the United States, 2008-2018: Rapid increases among young adults. J Psychiatr Res. 2020 Nov;130:441–446. eng. None declared. Epub 20200821. doi:10.1016/j.jpsychires.2020.08.014. Cited in: Pubmed; PMID 32905958.

16. Druet-Cabanac A, Azzi J, Lucchino M, Simon V, Offredo L, Briere JB, Hood S. Generalized anxiety disorder: epidemiology, burden, and comorbid depression. Curr Med Res Opin. 2025 Jun;41(6):1053–1064. eng. Epub 20250710. doi:10.1080/03007995.2025.2529974. Cited in: Pubmed; PMID 40611531.

17. Guimarães GO, D’Angelo F, Brouillette K, Souza LDM, da Silva RA, Mondin TC, Pedrotti Moreira F, Kapczinski F, de Azevedo Cardoso T, Jansen K. Incidence and risk factors for anxiety disorders in young adults: A population-based prospective cohort study. Encephale. 2023 Dec;49(6):572–576. eng. Epub 20221015. doi:10.1016/j.encep.2022.08.012. Cited in: Pubmed; PMID 36253174.

18. Tyrer P, Baldwin D. Generalised anxiety disorder. Lancet. 2006 Dec 16;368(9553):2156–66. eng. doi:10.1016/s0140-6736(06)69865-6. Cited in: Pubmed; PMID 17174708.

19. Pengpid S, Muanido A, Peltzer K. Prevalence and associated factors of undiagnosed depression and generalized anxiety disorder among a national sample of women and men in Mozambique in 2022-23. J Affect Disord. 2025 Jul 15;381:525–531. eng. Declaration of competing interest The authors have no conflicts of interest to declare. Epub 20250405. doi:10.1016/j.jad.2025.03.202. Cited in: Pubmed; PMID 40194633.

20. Leung P, Li SH, Graham BM. The relationship between repetitive negative thinking, sleep disturbance, and subjective fatigue in women with Generalized Anxiety Disorder. Br J Clin Psychol. 2022 Sep;61(3):666–679. eng. All authors declare no conflict of interest. Epub 20220127. doi:10.1111/bjc.12356. Cited in: Pubmed; PMID 35084773.

21. de Souza SCS, Campanini MZ, de Andrade SM, González AD, de Melo JM, Mesas AE. Watching television for more than two hours increases the likelihood of reporting poor sleep quality among Brazilian schoolteachers. Physiol Behav. 2017 Oct 1;179:105–109. eng. Epub 20170530. doi:10.1016/j.physbeh.2017.05.029. Cited in: Pubmed; PMID 28576672.

22. Werneck AO, Hoare E, Silva DR. Do TV viewing and frequency of ultra-processed food consumption share mediators in relation to adolescent anxiety-induced sleep disturbance? Public Health Nutr. 2021 Nov;24(16):5491–5497. eng. Epub 20210127. doi:10.1017/s1368980021000379. Cited in: Pubmed; PMID 33500011.

